# Prediction of COVID-19 infection risk using personal mobile location data only

**DOI:** 10.1101/2023.08.22.23294419

**Authors:** Ahreum Jang, Sungtae Kim, Hyeongwoo Baek, Hyejung Kim, Hae-Lee Park

## Abstract

Predicting an individual’s risk of infectious disease is a critical technology in infectious disease response. During the COVID-19 pandemic, identifying and isolating individuals at high risk of infection was an essential task for epidemic control. We introduce a new machine learning model that predicts the risk of COVID-19 infection using only individuals’ mobile cell tower location information. This model distinguishes the cell tower location information of an individual into residential and non-residential areas and calculates whether the cell tower locations overlapped with other individuals. It then generates various variables from the information of overlapping and predicts the possibility of COVID-19 infection using a machine learning algorithm. The predictive model we developed showed performance comparable to models using individual’s clinical information. This predictive model, which can be used to predict infections of diseases with asymptomatic infections such as COVID-19, has the advantage of supplementing the limitations of existing infectious disease prediction models that use symptoms and other information.

## Introduction

Beginning from early 2020, the COVID-19 pandemic caused significant human damage worldwide, and in the absence of treatments and vaccines for the newly emerged infectious disease, each country had no choice but to focus on epidemic prevention through non-pharmaceutical interventions[1]. Governments of each country implemented measures to reduce individual infection risk (wearing masks, social distancing, quarantine, lockdown, etc.) to prevent the spread of infectious diseases, and it was necessary to quickly lead individuals at high risk of infection to testing and treatment[2]. Implementing preventive measures for individuals at high risk of infection was one of the important response strategies in the early stages of the COVID-19 pandemic[3].

There are two typical ways to identify individuals at high risk of COVID-19 infection. One can identify high-risk individuals through the physical symptoms that appear when infected with COVID-19 and through the potential contact with an infected person. Firstly, screening high-risk individuals through several factors such as fever and respiratory symptoms, the main symptoms of COVID-19. Various machine learning and AI research to predict infection risk through these symptoms have been conducted, and there were models that showed a performance of up to 97.79% Accuracy[4–6]. However, this method has the disadvantage of missing asymptomatic infected individuals.

Another way to identify high-risk individuals is to trace those who have come into contact with an infected person. There have been attempts to utilize IT technology for this. Contact tracing mobile applications that track infected individuals and confirm whether they have come into contact with them were developed. These applications, which utilize widely used modern mobile devices and technologies such as Bluetooth and GPS (Global Positioning System), provided important information that could confirm individual locations and determine whether they had contact with infected people. Several open-source technologies emerged, and two companies providing mobile OS (Operation System), Apple and Google, even added technology to check and manage whether they came into contact with a COVID-19 infected individual in the mobile OS, and these digital contact tracing technologies are known to have been used in more than 46 countries[7,8]. These technologies are useful for both governments and individuals, but there is a limitation. These mobile applications must be installed and used by individuals themselves. In countries where it is not mandatory by the government, the actual usage rate of these applications was very low[9]. It’s a technology that is hard to see effects if there are few users.

South Korea took a different approach. Instead of contact tracing using mobile applications, they collected mobile cell tower location data from mobile carriers to find places visited by infected individuals and those who came into contact with them. It was possible due to South Korea’s high mobile usage rate and support of laws and systems. When a COVID-19 confirmed case occurred in South Korea, the goverment collected the cell tower location data for the 14 days prior to the COVID-19 confirmed patient’s PCR (Polymerase Chain Reaction) test date. This information was used as validation for epidemiological investigations conducted through interviews with the confirmed cases. Also, when a mass infection occurred with many confirmed cases, people who overlapped with the confirmed case at the cell tower location were deemed at risk of infection and were allowed to get tested. Individuals could not use this information directly and could get a COVID-19 PCR test if they were considered at high risk of infection based on information such as the visit places of infected individuals announced by the government[10]. Generally, it is not possible to know the exact location of an individual or whether they had contact with a specific person using cell tower location information[8]. Nevertheless, in South Korea, cell tower location information was used in epidemiological investigations of infected individuals and in analyzing infection hotspots[11].

The aim of this study is to develop a machine learning model that predicts an individual’s risk of COVID-19 infection using only cell tower location information. We conducted a study using COVID-19 test results and cell tower location information collected from a mobile application during the period of significant COVID-19 outbreak in South Korea. While it’s not possible to determine an individual’s exact location and whether they had contact with an infected individual with cell tower location information, we obtained results that we can predict an individual’s risk of COVID-19 infection using machine learning techniques. If we can address a few constraints discussed in the conclusion of this study, it can be used as a technology to prevent the spread of a pandemic by complementing the disadvantages of existing methods of finding high-risk infections in an infectious disease pandemic situation.

## Methods

### Data

All data were obtained from the SHINE mobile application. SHINE is an application that provides location-based COVID-19 outbreak information and COVID-19 coping strategies based on user-recorded information such as location, gender, age, COVID-19 test results, and vaccination data, etc. The app was launched on October 13, 2021, and operated until March 31, 2023, and was available for use through Apple’s AppStore and APK file installation(Android only) for individuals aged 14 and above (Fig 1). Upon registration on the mobile app, users consented online to the use of their de-identified device GPS data and personal information entered in the app for research purposes. Moreover, users who were subscribers of KT (Korea Telecom) also provided online consent to extract and utilize cell tower location data from KT’s network data infrastructure. Location data was stored only for the 14 days preceding the date when users recorded information such as PCR test results in the app. The COVID-19 PCR test results were uploaded directly to the application by the users themselves, and the application service administrators filtered out inaccurate information by comparing all uploaded test results – verified by documents issued by hospitals or screenshots of text messages with test results featuring the individual’s name – with the personal information entered during registration. Consequently, from the app’s launch to June 30, 2022, there were 43,270 total users, 21,046 users who registered PCR test results, and there were 17,678,028 cases of mobile cell tower location logs.

**Fig 1.**
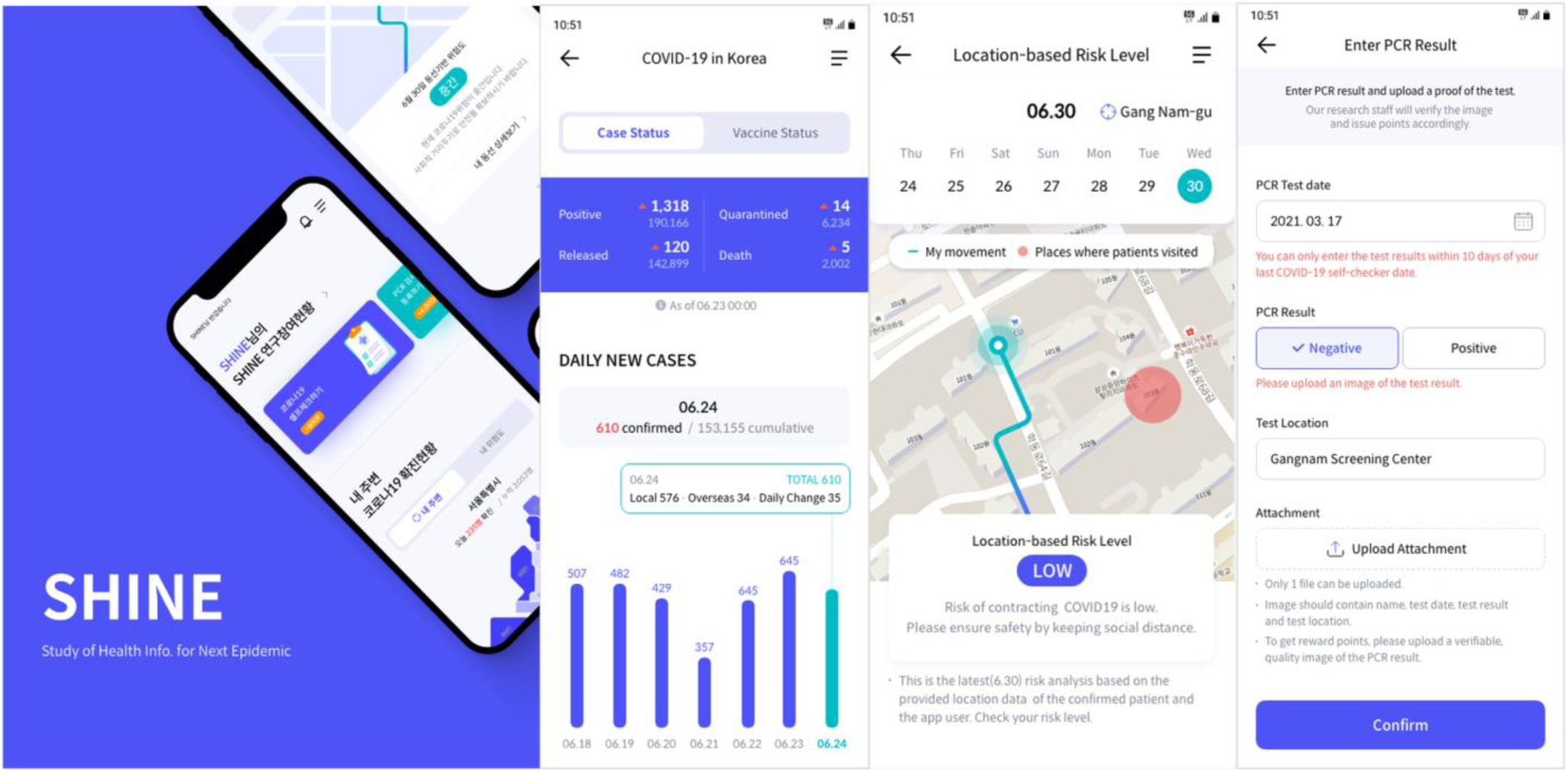
Screenshots of the SHINE mobile application. The language in the image has been translated into English.

This research received exemption from review by the Institutional Review Board of Sungkyunkwan University on November 11, 2022(IRB Number: SKKU 2022-11-014). Subsequently, we accessed location data, PCR test information, gender, and age data, which were collected retrospectively only from users who had consented for research purposes, from October 13, 2021, to June 30, 2022. Our access to this data began on November 28, 2022, and it ensuring all data were non-identifiable at the personal level.

The personal location information used in this study consists of mobile cell tower location data. This is data from regular communication between the subscriber’s device and the cell tower, containing information like timestamp, GPS location of the cell tower, and a unique identifier for subscribers[8]. This data has two limitations. The first is that the GPS location of the cell tower is not the precise location of the user but only indicating that the user is within the service radius of the cell tower. The second is that only information from KT subscribers can be used, and KT subscribers represent 31.3% of all mobile users in Korea[12]. These limitations require necessary assumptions and interpretations, which are discussed in the Discussion section.

On the other hand, the GPS information from the device collected in the mobile application was not used. Although the SHINE mobile application was set to collect the device’s GPS records in the mobile OS’s background, there were difficulties in continuously collecting reliable location records. Users often denied providing location information due to battery consumption from running the application in the background. Moreover, the mobile OS regularly sent location access approval or denial notifications to users about applications requesting device location information, leading to limited data tracking users’ locations continuously. However, we were able to obtain mobile telecom cell tower location data irrespective of the user’s mobile application usage, enabling us to know the reliable location of the user.

We extracted only KT users who could use cell tower location data among those who uploaded COVID-19 PCR test results on the mobile app. We further narrowed down the data to include only Seoul residents from January 1, 2022, to June 30, 2022. The reason for these limitations is that this period saw the largest outbreak of COVID-19 in Korea[13], and we had the most COVID-19 PCR test results data, and Seoul is the city with the highest population density in Korea[14]. There was a need to limit the region to areas with high population density to observe overlapping individual locations in the data. After considering the incubation period of COVID-19 and PCR test dates, we used PCR test results and cell tower location data of 837 individuals whose cell tower location data was collected for the seven days before the PCR test for modeling. The demographic information of these 837 individuals is as in Table 1.

**Table 1.** Demographic info. of Data.

|  |  | COVID-19 PCR test Result |  |
| --- | --- | --- | --- |
|  |  | Positive | Negative |
| Total, n(%) |  | 703 (84%) | 134 (16%) |
| Sex, n(%) | Male | 225 (26.9%) | 43 (5.1%) |
|  | Female | 478 (57.1%) | 91 (10.9%) |
| Age, n(%) | 10-19 | 40 (4.8%) | 0 (0.0%) |
|  | 20-29 | 232 (27.7%) | 39 (4.4%) |
|  | 30-39 | 230 (27.5%) | 43 (5.1%) |
|  | 40-49 | 116 (13.9%) | 35 (4.2%) |
|  | 50-59 | 57 (6.8%) | 14 (1.7%) |
|  | 60-69 | 20 (2.4%) | 3 (0.4%) |
|  | 70-79 | 7 (0.8%) | 2 (0.2%) |
|  | 80-89 | 1 (0.1%) | 0 (0.0%) |

### Data preprocessing

First, we distinguished whether the cell tower location from each individual’s seven-day location data was a residential area or a non-residential area. We designated the cell tower location that appeared most frequently from 10 pm to 7 am the next day for each individual as the residential cell tower location. We then added a variable to each individual’s cell tower location record to distinguish whether it was a residential or non-residential area, anticipating that the infection characteristics would vary depending on this classification. We also extracted cell tower location records only from 9 AM to 10 PM. To assess the possibility of contact with others, as shown in Fig 2, for each individual’s cell tower location record, we checked the overlapping time with others at the same cell tower. If the overlap was more than 10 seconds, we marked it as an overlapping. The overlap criterion of 10 seconds was selected as it provided the largest difference in COVID-19 infection rates between user groups with at least one overlapping record and those without any overlapping records (Fig 3). Furthermore, in checking the overlap time at the same cell tower with others, we did not refer to the others’ PCR results information. This is because the others’ COVID-19 infection status cannot be confirmed when predicting an individual’s COVID-19 infection. The overlapping time information at cell tower locations for each individual over seven days was summarized into six types of information by distinguishing the type of cell tower location (residential or outing area), and the characteristics of all predictor variables used for modeling are as in Table 2. And all predictor variables were transformed using the natural logarithm to reduce data skewness.

**Fig 2.**
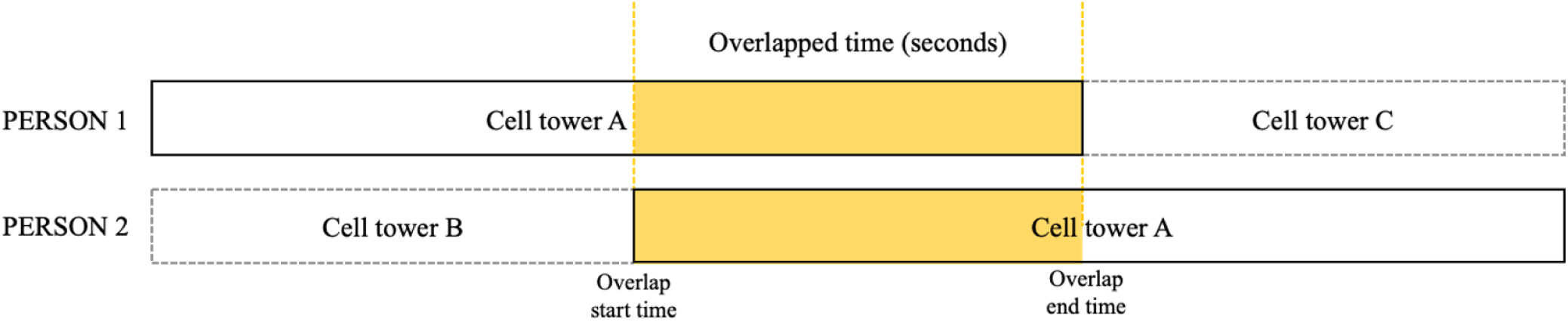
Method for calculating overlaps. In each individual’s location records, it was counted as an overlap if they were located at the same cell tower as another person for 1 second or more.

**Fig 3.**
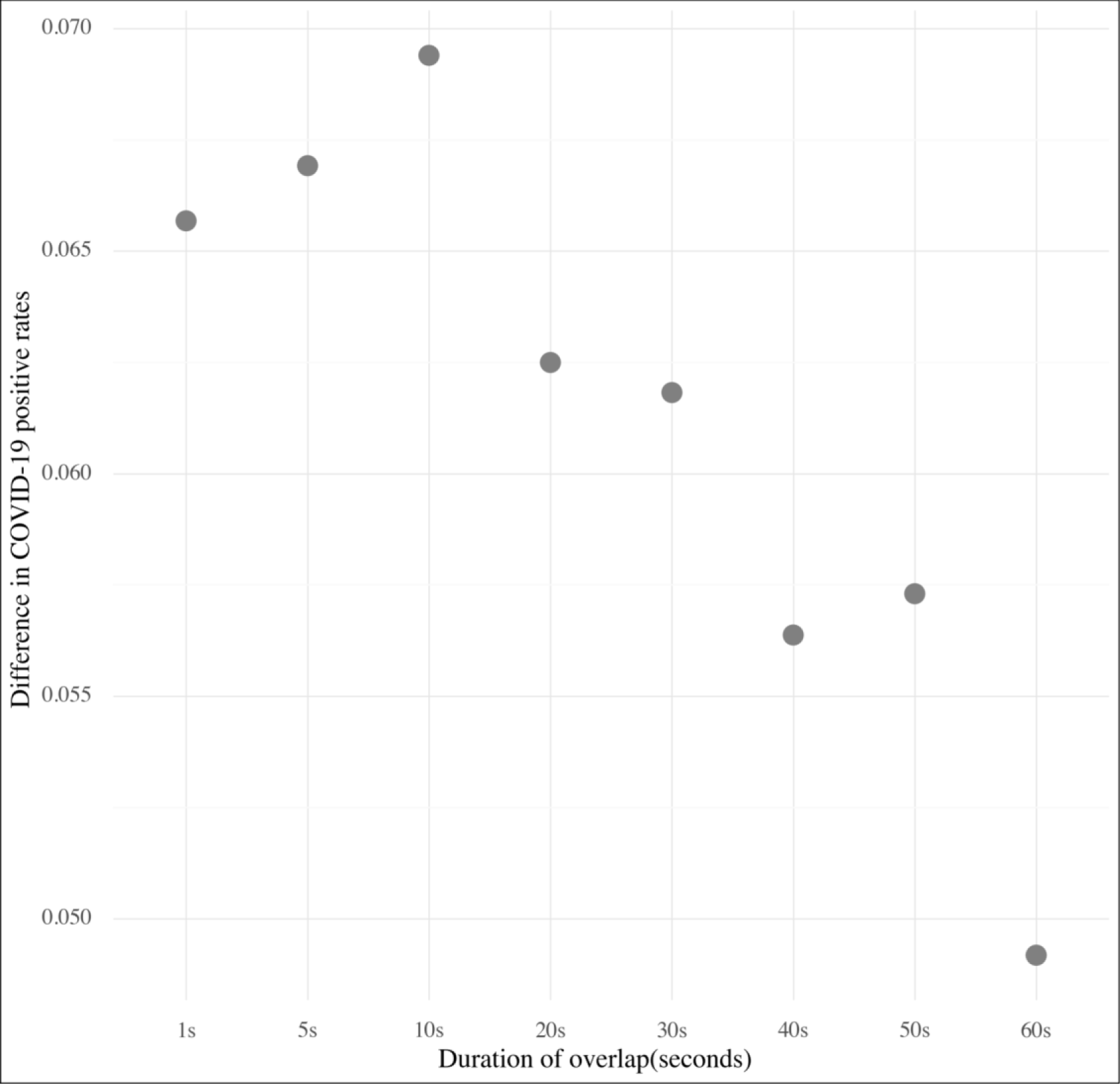
Difference in COVID-19 positive rates based on the duration of overlap (in seconds). Difference in COVID-19 positive rates by comparing user groups with overlapping records and those without, based on each duration of overlap.

**Table 2.** Characteristics of features.

| Variables |  | n | Mean | SD<sup>a</sup> |
| --- | --- | --- | --- | --- |
| Number of Overlaps | Outside | 837 | 38.62 | ±126.01 |
|  | Residence | 837 | 15.01 | ±63.71 |
| Number of Overlapped people | Outside | 837 | 1.4 | ±1.46 |
|  | Residence | 837 | 0.37 | ±0.64 |
| Total Overlapped Time<sup>b</sup>(seconds) | Outside | 837 | 888.98 | ±2956.47 |
|  | Residence | 837 | 302.01 | ±1353.17 |
| Max Overlapped Time<sup>c</sup>(seconds) | Outside | 837 | 75.59 | ±199.36 |
|  | Residence | 837 | 25.32 | ±77.47 |
| Average Overlapped Time<sup>d</sup>(seconds) | Outside | 837 | 0.33 | ±0.86 |
|  | Residence | 837 | 0.27 | ±0.91 |
| Minimum Overlapped Time<sup>e</sup>(seconds) | Outside | 837 | 9.16 | ±12.59 |
|  | Residence | 837 | 3.64 | ±11.13 |
| Number of overlapped locations(outside) |  | 837 | 4.17 | ±11.65 |

<sup>a</sup>SD = Standard Deviation
<sup>b</sup>Total Overlapped Time = Sum of all time overlapped with others over 7 days.
<sup>c</sup>Max Overlapped Time = Max duration of time overlapped with others in a single event over 7 days.
<sup>d</sup>Average Overlapped Time = Average duration of time overlapped with others per event over 7 days.
<sup>e</sup>Minimum Overlapped Time = Minimum duration of time overlapped with others per event over 7 days.

### Development of model for prediction

To train the model, the data was divided into a Training data set (585/837, 70%) and Test data set (252/837, 30%). It was ensured that the ratio of COVID-19 PCR test results was maintained during this division. To enhance the model’s generalization performance, records that corresponded to outliers were removed from the Training data set prior to model training. Outliers were determined by calculating the IQR (Interquartile Range) for each variable; values smaller than 1.5 times the first quartile or larger than 1.5 times the third quartile were considered as outliers. Subsequently, data with negative COVID-19 PCR test results was subjected to Random over-sampling, so the ratio of positive to negative results for the target variable, the COVID-19 PCR test result, became 1:1. We experimented with machine learning models like Logistic Regression, a binary classification model, XGBoost Classifier, and Random Forest Classifier. These models are frequently used for classification and were used in this study to develop a COVID-19 infection prediction model. Python (version 3.8.10)’s scikit-learn (version 1.0.2) library’s GridSearchCV was used to train the Training data set using 10-Fold Cross Validation. The entire process from data preprocessing to machine learning model training is as described in Fig 4.

**Fig 4.**
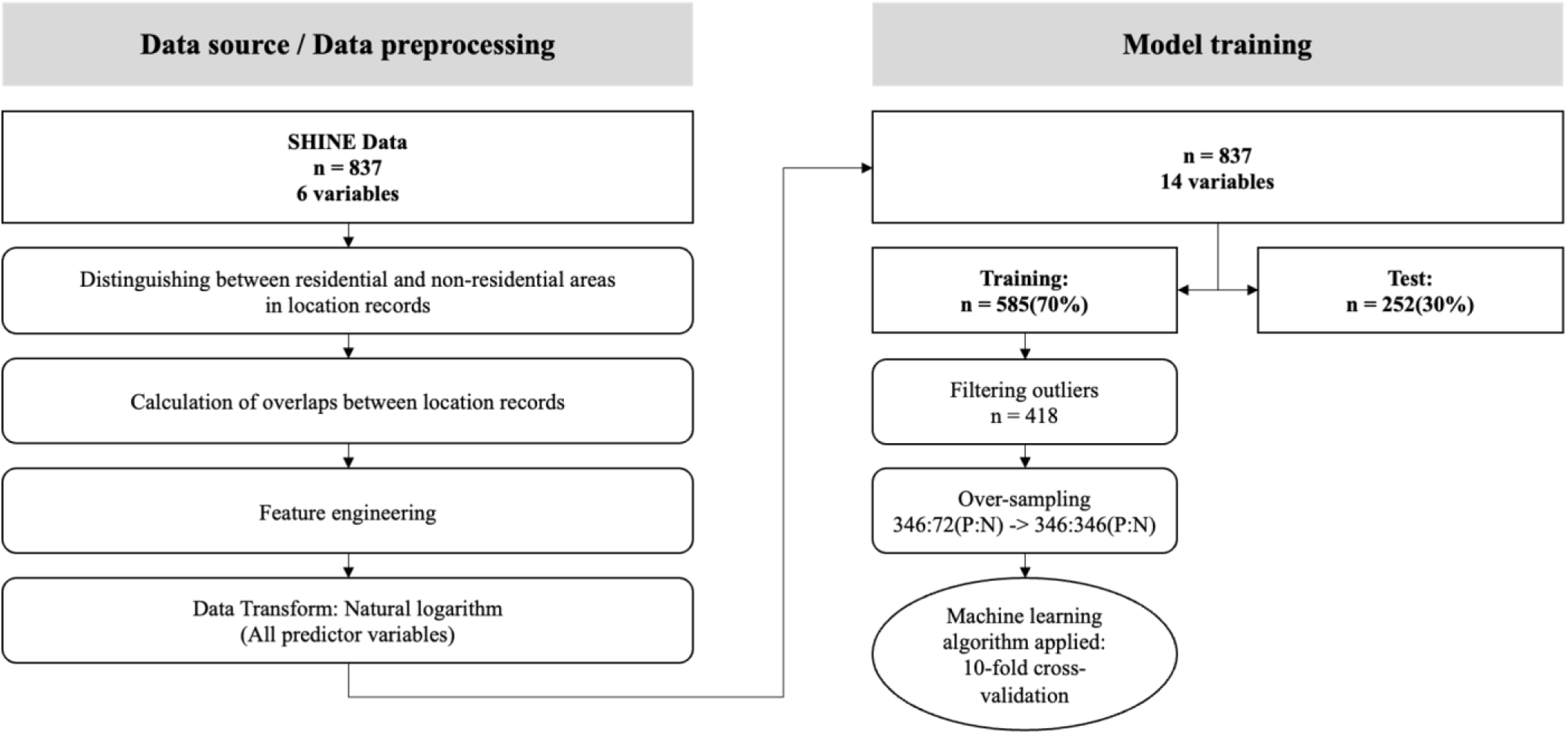
Workflow of analysis.

## Results

Table 3 shows the performance results measured in the Test data set with five metrics for the three trained models. Logistic regression exhibited the lowest performance across all metrics, while XGBoost showed the highest Accuracy and Sensitivity. In contrast, the Random Forest model demonstrated high Specificity and Precision, outperforming both the Logistic Regression and XGBoost models in terms of Specificity. There was no significant difference in AUC(Area Under the Curve) performance between Random Forest and XGBoost, but among the models tested, Random Forest had the best AUC. The ROCs (Receiver Operating Characteristic curves) for the three models are shown in Fig 5.

**Fig 5.**
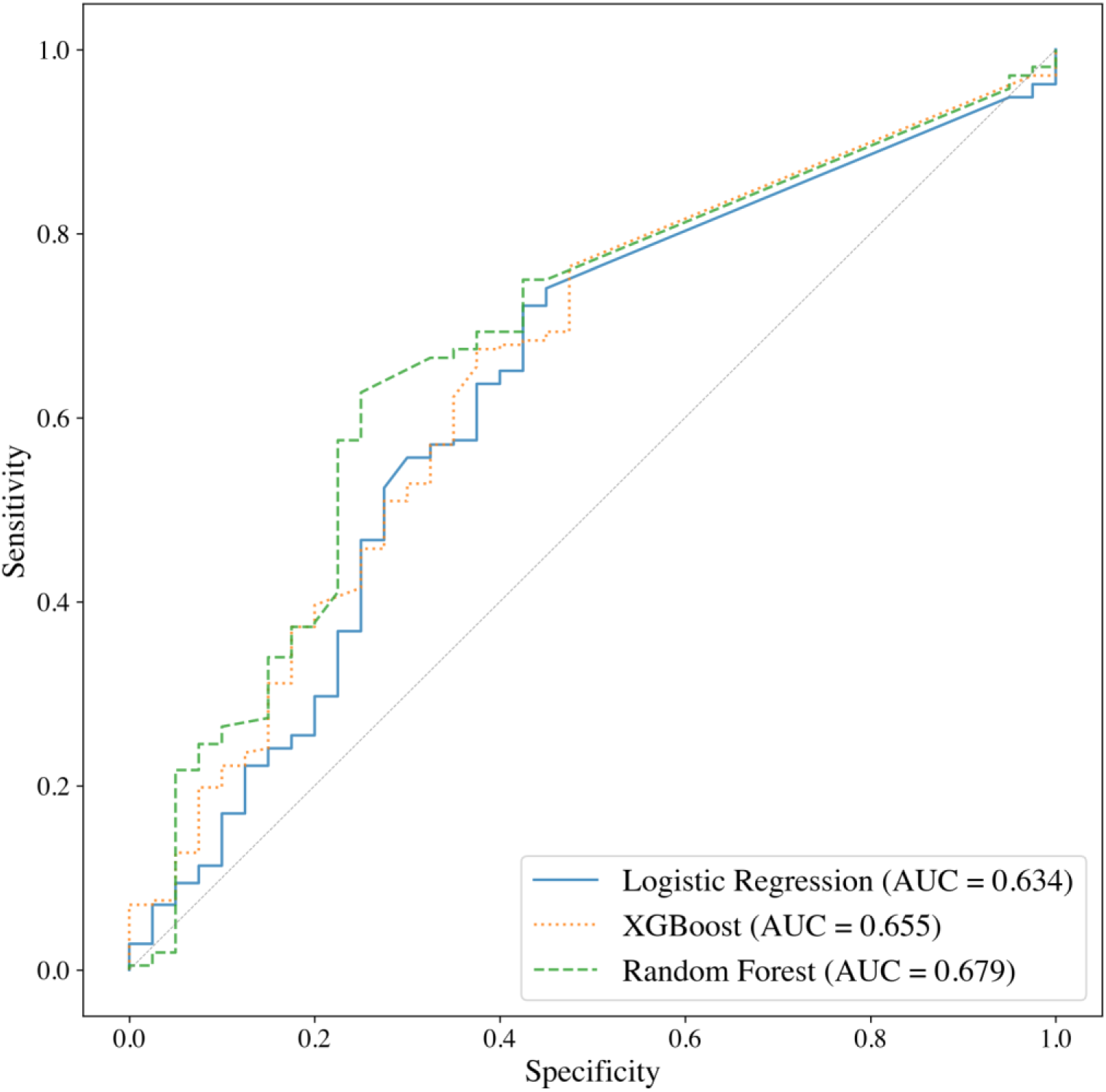
Receiver operating characteristic curves with corresponding AUC values. AUC values for each model are also presented in Table 2.

**Table 3.**
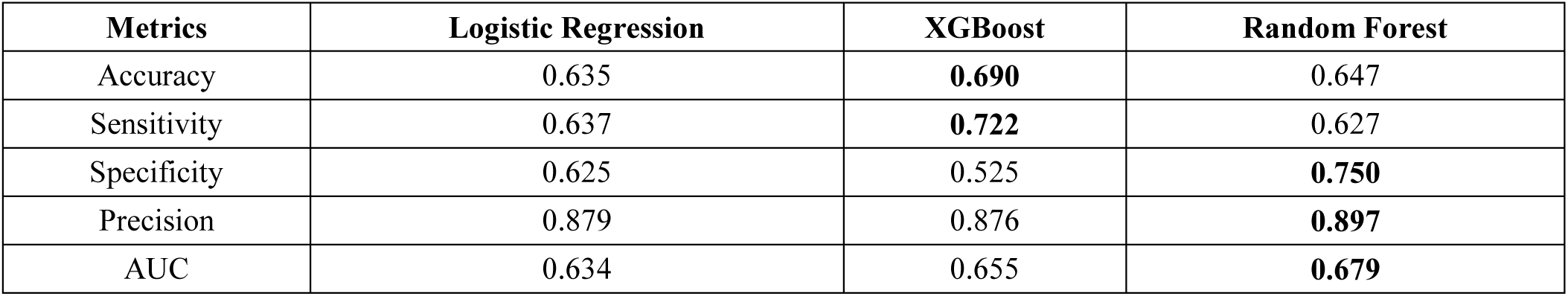
Performance of machine-learning algorithms and logistic regression.

The rank of importance of predictor variables in the Random Forest model, which had the highest AUC, is shown in Fig 6. The variable with the highest importance was the Total Overlapped Time in non-residential areas. Also, variables related to non-residential areas had higher importance than those related to residential areas. This finding suggests that activities conducted outside residential areas can provide significant insights for predicting COVID-19 infection.

**Fig 6.**
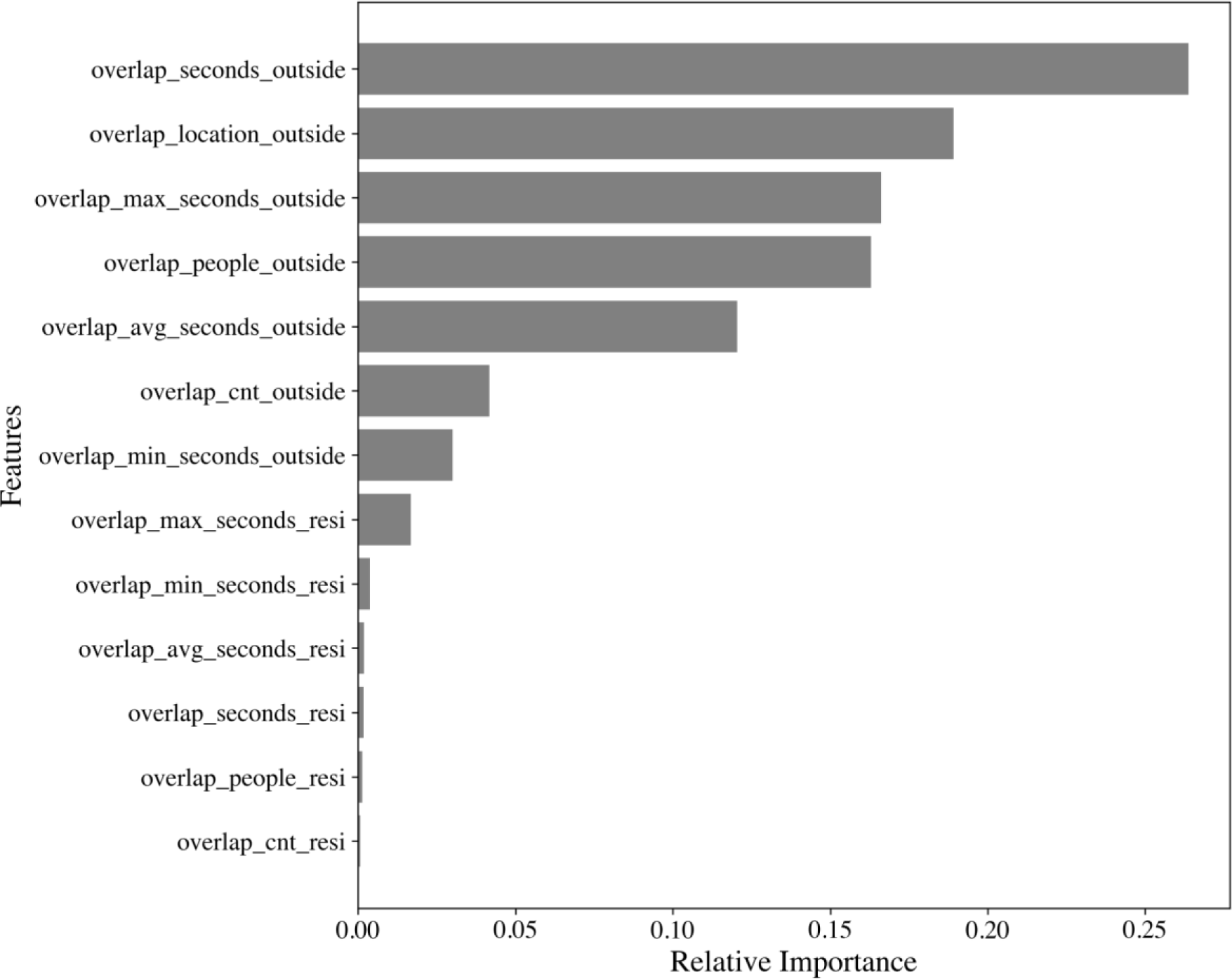
Variable importance plots of COVID-19 PCR result predictors for Random forest.

Table 4 summarizes the algorithms, data, sample sizes, and performance of existing studies related to COVID-19 infection prediction. The six studies we reviewed all primarily utilized individual demographics and clinical information, and performance ranged from 0.689 to 0.98 based on AUC (limited to studies that disclosed AUC). Compared to these studies, our developed model’s performance was relatively low. However, despite our model utilizing only individual location records for predictions, its AUC did not significantly differ from that of models incorporating individual symptoms [15]. When comparing the results of our Random Forest model with the results of this study, our model showed lower AUC and Sensitivity, but higher Specificity and Precision.

**Table 4.** Comparison with the Prediction model using symptoms.

| Reference | ML/AI methods | Types of Data | Sample | Performance |
| --- | --- | --- | --- | --- |
| [16] | Support Vector Machine | Clinical, laboratory features, Demographics | 556 | Accuracy: 77.5%<br>Specificity:78.4% AUC: 0.98(testing dataset) |
| [17] | Random forest | Clinical, Demographics | 253 | Accuracy: 95.95% Specificity: 96.95% |
| [18] | Logistic regression, Random Forest, Decision tree, Linear SVM, Naive Bayes, Gradient boosting classifier | Clinical, Demographics | 5,434 | Accuracy: 97.79%, Sensitivity: 0.99, Precision 0.97 |
| [15] | Logistic Regression | Clinical | 378 | AUC: 0.6891, Specificity: 58.6%, Sensitivity: 64.7%, Precision: 43.1% |
| [19] | Random Forest | Clinical, Demographics | 1,653 | AUC: 0.788, recall: 0.799, FPR: 0.38 |
| [20] | Logistic Regression (LASSO) | Clinical, Demographics | 143,531 | AUC: 0.78 |

## Discussion

We have developed a prediction model that addresses the limitations of existing methods in predicting high-risk individuals for COVID-19 infection. We did this by creating a variable that represents the possibility of contact with others using only an individual’s cell tower location information, and then developing an infection risk prediction model using machine learning algorithms. Though our model showed lower performance compared to the infection prediction model using symptom information, our results indicate that an individual’s COVID-19 infection status can be predicted to a certain degree without relying on explicit symptoms or contact tracing information. This could potentially address the limitations of existing studies, which struggle to predict the infection risks of asymptomatic carriers, and of mobile contact tracing applications with a low user base. Moreover, as shown in Fig 6, our study supports the general characteristic of infectious diseases that a higher possibility of contact with others leads to a higher risk of infection.

Another point worth discussing is the overlap information we used for model development. We created a variable for overlap information if the cell tower locations of each individual overlapped with others for more than 10 seconds. However, an overlap of cell tower locations does not necessarily imply direct contact between two individuals. Furthermore, the overlap of cell tower locations for 10 seconds does not confirm the transmission of COVID-19. The data we used represents only a very small part of all Korean citizens’ cell tower location records, and it is not possible to conclude that an individual was infected with COVID-19 from the people they overlapped locations with based on this information alone. We believe this data does not suggest that an individual contracted COVID-19 from someone with whom they overlapped locations, but rather indirectly indicates they were in a location with a high risk of COVID-19 infection. In our sample of 837 people, the places where locations overlapped are likely to have seen many more overlapping individuals, thus increasing potential contact points. As shown in Fig 6, we believe the risk of COVID-19 infection increases when an individual frequently overlaps cell tower locations outside their residence, thereby increasing potential contact with others.

In order to apply the findings of this research, the following prerequisites need to be fulfilled. The government or the institution intending to utilize these research results should be able to collect cell tower location information from mobile carriers. The Korean government was able to collect cell tower location information of confirmed cases without the individual’s consent through legal procedures. While it is not necessary for many users to install and use the contact tracing app, the system and technology should allow for the collection of individual cell tower location information.

Additionally, we foresee the following further research to enhance our findings. First, important conclusions were derived from a relatively small sample of 837 people within a limited period and geographic scope. However, these results emphasize the need for further research based on a larger dataset. If the study is expanded to include a significantly larger number of people, more substantial conclusions could be drawn. Second, it would be beneficial to investigate whether the risk of infection from diseases other than COVID-19 can also be predicted using cell tower location information. For infectious diseases with varying characteristics, the accuracy of infection prediction using cell tower location information may vary, and this is important consideration. Lastly, we believe that by incorporating individual symptom information, a more accurate infectious disease prediction model could be developed.

## Data Availability

All data produced are not available. Raw data cannot be shared publicly because of the sensitive nature of the personal information collected, which includes location data and COVID-19 test results. According to South Korean law, unless individual consent is specifically obtained from the data subject, this information cannot be transferred to third parties.

## Acknowledgements

This research is based on the researchs “A Next Generation Surveillance Study for Epidemic Preparedness(INV-006404)” which was funded by the Bill & Melinda Gates Foundation, and “COVID-19 Self-Risk Evaluation with Digital Contact Tracing(RF-TAA-2020-D07)” which was funded by the RIGHT Foundation. The findings and conclusions contained within are those of the authors and do not necessarily reflect positions or policies of the Bill & Melinda Gates Foundation.

## Supporting information

**S1 Table.** Description of software packages, methods and tuning parameters for model development.

| Algorithm | Package / Method | Parameters | Final chosen model |
| --- | --- | --- | --- |
| Logistic Regression | scikit-learn / LogisticRegressionCV | solver='liblinear' max_iter=1000 |  |
| XGBoost | xgboost / XGBClassifier | n_estimators=range(200, 310, 10)<br>learning_rate=[0.1] max_depth=range(8, 11)<br>alpha=[0, 0.1] gamma=[0, 0.1] lambda=[0, 0.1] subsample=[0.4, 0.5, 0.6, 0.7, 0.8] | alpha=0 colsample_bytree=0.7<br>gamma=0.1 lambda=0<br>max_depth=8 |
|  |  | colsample_bytree=[0.4, 0.5, 0.6, 0.7, 0.8]<br>min_child_weight=range(1, 9)<br>objective=['binary:logistic'] | n_estimators=260<br>subsample=0.8 |
| Random forest | scikit-learn / RandomForestClassifier | n_estimators=range(200, 300, 10)<br>max_depth=range(3, 11)<br>max_features=range(4, 13)<br>max_leaf_nodes=range(4, 13)<br>min_samples_leaf=range(5, 11)<br>min_samples_split:range(5, 11) | max_depth=10<br>max_features=12<br>max_leaf_nodes=13<br>min_samples_leaf=7<br>min_samples_split=5<br>n_estimators=220<br>random_state=42 |

